# Biomonitoring Fumonisin B1 and Zearalenone to improve public food safety for female cereal value chain workers in Ghana

**DOI:** 10.1101/2025.02.23.25322751

**Authors:** N.K-A. Quartey, J. Kumi, H.E. Lutterodt, G.M. Ankar-Brewoo, J. Ampofo, W.O. Ellis, I.W. Ofosu

## Abstract

Monitoring biomarkers offers more reliable results, compared to typical mycotoxin exposure estimation approaches that employ their prevalence in food in combination with population food intake data. Free forms of fumonisins B1 and B2 (FB1+ FB2), and free zearalenone (ZEN) together with three of its enteric metabolites have been identified as fitting indicators for exposures to the parent mycotoxins. This study investigated the similarities in regional and age-related mycotoxin exposures among sampled females employed in the cereal value chain in Ghana. After obtaining ethical clearance to conduct the study, spot urine samples were collected from females in cereal growing communities in The Upper West, Northern, Ashanti and Greater Accra regions of Ghana. The collected bio-samples were processed appropriately and LC-MS/MS chromatographic methods used to determine the presence and levels of FB1, FB2, ZEN, α-ZEL, β-ZEL and ZEN-14, glucuronide. The results indicate that while probable daily intake of the mycotoxins decreased with age, being higher for the 10-19 age groups, exposures were higher in the 55+ age group, consistent with age-related accumulation. The creatinine adjusted mycotoxin levels also presented a snapshot of renal function across the four regions being generally homogenous for participants in the Ashanti region. Wide variations in creatinine concentrations were observed for the Upper West and Northern regions and the spread was heterogenous for Greater Accra. The insight into age-related mycotoxin exposures agrees with previous findings of significant links between FB1 exposures and adverse health outcomes. ZEN exposures were higher in the 10-19 age group suggesting the potential for younger age groups to face significant risks due to exposure to the mycotoxin. The findings from this biomonitoring exercise prompts heightened public health interventions, guided by strategies that will adequately address the age-related risk of mycotoxin exposures in different populations.

## 1.0 Introduction

Biomonitoring of biological markers has recently been shown to be more reliable for determining biohazard exposures than traditional risk assessment methods (1,2). This process involves measuring biomarkers in biological fluids such as urine, blood, and sometimes stool to assess human exposure to environmental and dietary hazards (3). Biomonitoring exercises provide invaluable information about the internal doses of chemical hazards, reflecting the accurate hazard concentrations individuals have been exposed to through diet or tactile contact (4). The findings from biomarker detection in human bio-samples can help establish linkages between chronic exposures to environmental pollutants and adverse health outcomes. This information can further help estimate disease burdens attributable to these biohazards (2,5,6), aiding in addressing public health concerns and shaping health policies.

Fumonisin B1 (FB1) and Zearalenone (ZEN), are two mycotoxins produced primarily by *Fusarium spp.* fungi and are known for impacting human health due to their pervasive and frequent occurrence (5,7). FumonisinB1, a Group 2B carcinogen (8), is produced primarily by *Fusarium verticillioides, Fusarium proliferatum,* and *Fusarium nygama* (9). The non-carcinogenic Group 3 zearalenone is produced by *F. verticillioides, F. graminareum, F. culmorum,* and *F. equiseti* (8,9). According to International Agency for Research on Cancer (IARC) classification, Group 2B substances are possibly carcinogenic to humans while Group 3 substances have not been classified as to their carcinogenicity due to inadequate evidence (10).

While FB1 has been shown to cause health issues ranging from liver to kidney damage, ZEN exerts intense estrogenic activity, leading to toxicity in female reproductive and developmental systems (11). The frequency and co-occurrence of these two mycotoxins in cereals and their products place them as priorities (12,13) when considering biohazards with the potential to trigger adverse health effects on exposure. Populations depending heavily on cereals are constantly at risk of exposure to these two mycotoxins, and chronic exposures are rife in communities consuming maize as a staple food (11,14). Mitigation measures remain lax and unconcerted without an accurate representation of exposure levels (15).

Using the biomonitoring approach to directly measure specific metabolic biomarkers of the mycotoxins, a true reflection of dietary exposures, can be achieved (16). FB1 is the most toxicologically potent B-type fumonisin and is highly stable (9,17), during incubation although it can be hydrolysed in human kidney and liver cells. Its low metabolization lends to its stability *in vivo*, making it possible for hydrolysed (HFB1) and free forms of FB1 to be detected in bodily excretions (9). Human biomonitoring studies using urine have focused on measuring free FB1 concentrations, as it is the most prevalent of the FB mycotoxins, followed by FB2 and then FB3. A statistically significant (p<0.05) correlation exists between dietary FB and urinary FB1 levels based on Pearson product correlation (18).

Zearalenone has been found to cause significant fertility problems in animal models (9). Unlike the stable FB1, the toxicokinetics of ZEN indicate that it is extensively and quickly absorbed from the gastrointestinal tract and undergoes Phase I and Phase II reactions to yield metabolites (19). ZEN is suspected to be a key etiologic agent in foetus and infant intoxication, consequently manifesting in breast enlargement, pubarche, and premature thelarche (9). Pubarche, the earliest sign of puberty, is the first appearance of pubic hair, and thelarche defines the beginning of breast development in girls at the start of puberty (20,21). These outcomes are as a result of ZEN actively competing with estrogen for binding sites, earning the mycotoxin an association with females and tagged as a mycoestrogen (22,23). Notwithstanding its Group 3 classification by the IARC, ZEN stimulates the growth of human breast cancer cells (17). ZEN is reduced in vivo to its primary metabolites α-zearalenone (α-ZEL) and β-zearalenone (β-ZEL) in a reversible process in liver microsomes (24). Although as many as 14 glucuronides of ZEN have been identified in animal models, only three have been detected in human urine (9); ZEN-16-glucuronide (ZEN-16-GlcA), ZEN-14-glucuronide (ZEN-14-GlcA), and ZEN-14, 16-glucuronide (ZEN-14, 16-GlcA).

Despite mycotoxins’ known adverse health effects, only traditional dietary risk assessment methods remain prevalent for evaluating exposures in Ghanaian communities (25,26). These methods, relying on food recall and frequency data, often face significant uncertainties, making it challenging to estimate exposures accurately (14). Consequently, there is a gap in accurately assessing the internal doses and chronic exposure levels of mycotoxins among the population (27). Women constitute about 40% of the global agricultural workforce (28) and play crucial roles in the food value chain from farm to fork. In Ghana, approximately 48% of labour in agriculture are females and 70% of food production is also handled by females (29). Women employed in farming have been found to experience higher burnout than male counterparts (30) making them more vulnerable to ailments (31) and chronic diseases. Moreover, acute and chronic exposures to chemical hazards for females in the cereal value chain warrants special attention due to the carcinogenic and teratogenic nature of some chemicals, including mycotoxins (32). The vulnerability of females in the cereal production value chain to adverse health outcomes necessitate constant monitoring in view of potential mycotoxin exposures. In Ghana, communities that heavily rely on cereals, mainly maize, are frequently and significantly exposed to co-occurring FB1 and ZEN (14). The underestimation of exposure levels results in insufficient mitigation measures, leading to severe public health risks, particularly in terms of liver, kidney, and reproductive health issues for females. This leads to broader implications for public health, sustainability, and economic development (17). Addressing these challenges is crucial for aligning with the Sustainable Development Goals (SDGs), specifically Good Health and Well-being (SDG 3) and Zero Hunger (SDG 2)(3). The primary objective of this study was to conduct a biomonitoring exercise to accurately determine the presence and concentrations of urinary biomarkers of Fumonisin B1 (FB1) and Zearalenone (ZEN) among females in selected cereal-growing communities in Ghana. The realistic and timely evaluation of chronic mycotoxin exposures will ensure determination of the probable daily intake of these mycotoxins based on internal exposures.

Consequently, the data will accurately interpret epidemiological health outcomes and inform effective preventive and control measures for food mycotoxin contamination (27).

## 2.0 Materials and Methods

The study was approved and conducted per ethical guidelines governing research. All participants were informed about the nature of the study, and written consent was obtained before their inclusion. For participants below 18 years of age, verbal consent was obtained in addition to a signed informed consent form from a guardian or parent. All participants were informed of their right to withdraw from the study at any time without providing a reason. No medical examinations were conducted in this study.

### 2.1 Study Area

The biomonitoring exercise was carried out in randomly selected communities across four regions in Ghana (Figure 1) known for cereal production. The study areas are spread across the Greater Accra, Ashanti, Northern, and Upper West regions. These regions were chosen due to their significant contribution to cereal production and the high proportion of female residents involved in the cereal value chain. Major trading towns in these regions, especially on market days, rely on the selected communities for cereal produce. Crops such as maize, millet, rice, and sorghum are cultivated to varying extents, with the maize value chain being particularly prominent, followed closely by rice.

**Figure 1:**
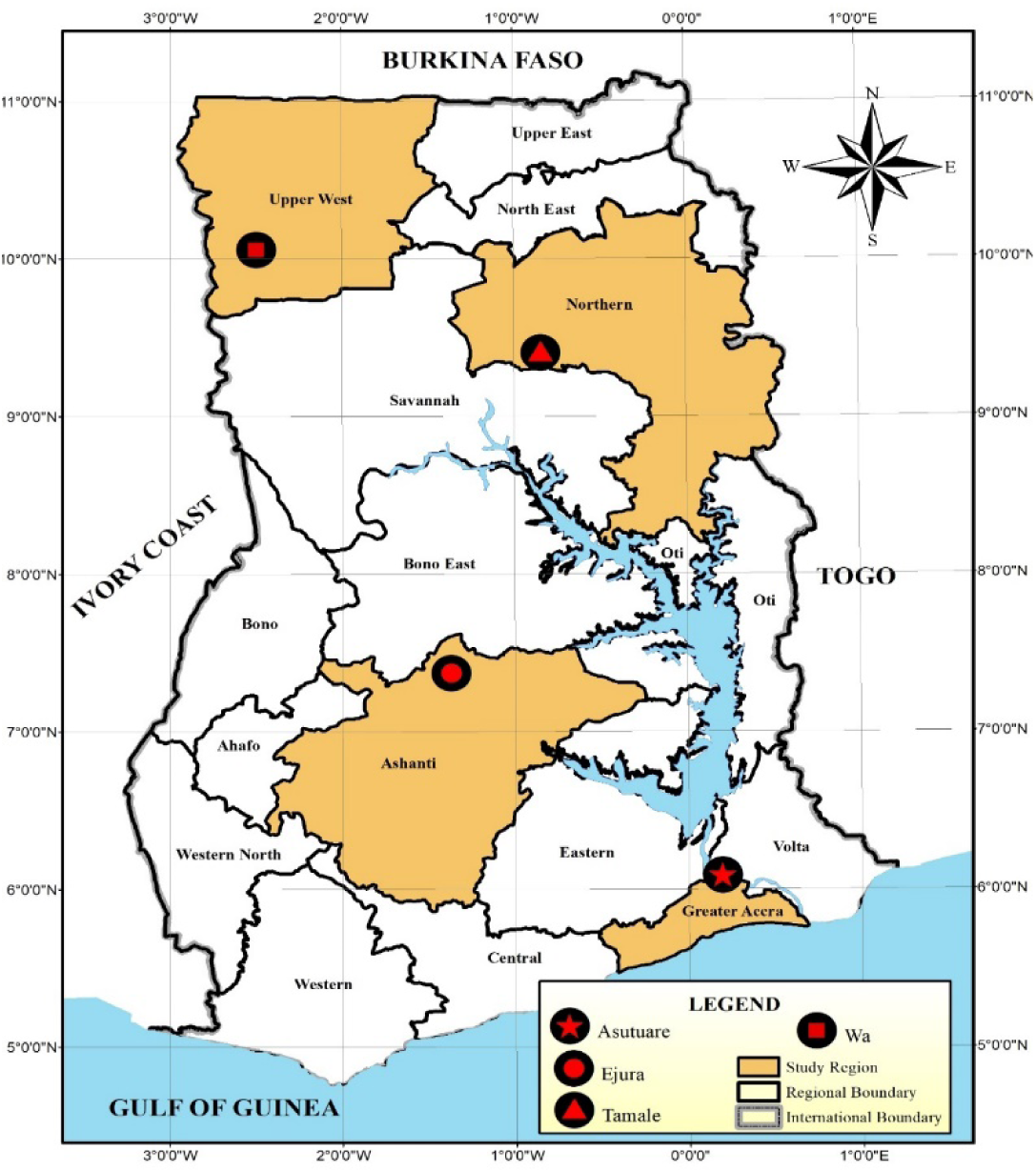
Sampling sites of the biomonitoring exercise.

Biomonitoring activities were conducted across key locations in the four regions. These included Asutsuare in the Greater Accra Region, a farming town renowned for its fish and cereal production, primarily rice. Other locations included Wa in the Upper West Region, a significant maize and rice cultivation centre, and Tamale and Kumbuyili in the Northern Region, with Kumbuyili being a dispersed settlement along the Tamale-Kumbungu Road. Within the Ejura Municipality of the Ashanti Region, biomonitoring was conducted at six distinct sites: Aframso, Anyinasu, Babaso, Frante, Homako, and Nokwareasa. Aframso, Babaso, Homako, and Nokwarease are within a 10-mile radius of Ejura, while Anyinasu and Frante fall within a 20-mile radius.

### 2.2 Subjects and Sampling

Starting March 9, 2022 to February, 15 2023, spot urine samples were collected from willing females residing in cereal-growing communities across Ghana, adhering to an approved study protocol. Female participants (≥ 10 y) were prioritized due to the mycoestrogenic properties of Fusarium species’ mycotoxins. The required sample size was determined (33) using Equations 1a and 1b:

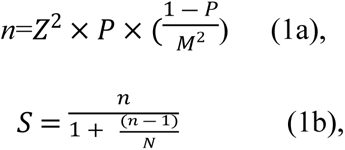

Where *n* is the sample size of an infinite population, Z is the z-score corresponding to a 95% confidence interval, M is a 5% margin of error, P is a 50% probability of response, S is the adjusted sample size, and N is the known population size of the study area.

Regional Health Officers (RHO) in the study sites were contacted to facilitate sample collection. Community Health Nurses (CHN) subsequently facilitated community entry and participant recruitment. Sampling employed a mixed approach, combining random cluster sampling with snowball sampling, where some participants joined voluntarily while others were encouraged to participate upon observing the involvement of their peers. Sample collection was conducted at the community health facility under the supervision of the CHN and with authorization from the RHO. All participants provided informed consent and completed a detailed questionnaire encompassing demographic information and a food frequency questionnaire. Exclusion criteria included all males, female children under 10 years of age, individuals seeking medical attention for unrelated ailments, and pregnant women in their second and third trimesters. A simple pregnancy test kit was utilized to confirm pregnancy for this exclusion criterion.

### 2.3 Urine sample collection and preparation

Spot urine samples were collected from participants using sterile 50 mL urine sample cups. Samples were maintained under cold conditions using ice packs within field sample boxes before transportation to the laboratory. Upon arrival, samples were processed for long-term storage at - 20 °C within 6 h of collection.

### 2.4 Reagents and Chemicals

HPLC-grade acetonitrile and methanol were procured from Sigma-Aldrich (St. Louis, MO, USA). Ammonium acetate and formic acid were also obtained from Sigma-Aldrich. Distilled water was produced using a Milli-Q plus apparatus (18 Ω, Millipore, Billerica, MA, USA). Analytical standards for Fumonisin B1 (FB1), Zearalenone (ZEN), α-Zearalenol (α-ZEL), β-Zearalenol (β-ZEL), and internal standard C18-zearalenone (U-13C18 ZEN) were acquired from Romer Labs Diagnostic GmBH (Tullin, Austria). The mycotoxin conjugate, Zearalenone-14-glucuronide (ZEN-14-GlcA), was obtained from Scientific Laboratory Supplies (Nottingham, UK).

### 2.5 Biomarker Analysis

The extraction and purification of urinary mycotoxins (FB1, ZEN, and their metabolites such as α-ZEL, β-ZEL, and ZEN-14-GlcA) were performed following previously established protocols (Fan *et al*., 2023; Föllmann *et al*., 2016; Heyndrickx *et al*., 2015; Huang *et al.*, 2021) with minor modifications. Urine samples were equilibrated at room temperature for 30 min and then vortexed for 30 s. A 1 mL aliquot was centrifuged at 9,800×g for 10 min to precipitate salts and proteins. A dilution solvent comprising distilled water, methanol, and 1% formic acid (94/5/1, v/v/v) was prepared. For the “dilute and shoot” method, 500 µL of dilution solvent was added to 500 µL of urine sample, filtered through a 0.22 µm syringe filter, and analysed using a 6420 Triple Quad LC/MS (Agilent Technologies, Santa Clara, CA, USA). Detailed chromatographic and MS parameters are provided in Supplementary S1, S2 and S3 of the Supplementary Material. The analytical methods underwent rigorous validation to assess linearity, sensitivity, and reproducibility, adhering to the guidelines outlined in Commission Decisions 2002/657/EC and 2006/401/EC (36).

### 2.6 Urine creatinine analysis

Biomarkers can be quantified directly in urine samples, providing raw, non-adjusted concentrations. However, these levels are influenced by factors such as hydration, diet, and sample collection time, thus hindering accurate inter-individual or inter-group comparisons (37). To address this challenge, creatinine-adjusted biomarker levels are often determined. The procedure involves dividing the raw biomarker concentration by the creatinine concentration in the same urine sample. Creatinine (Cr), a muscle breakdown waste product, is a normalization factor which accounts for urine concentration variations, improving biomarker comparison reliability and exposure assessment. Thus normalizing mycotoxin concentrations based on urinary creatinine (Cr) levels (µg/g Cr) (35,38).

Urinary creatinine concentrations were determined using a slightly modified spectrophotometric method (39). Briefly, 3.5 mM picric acid was reacted with 1000 mM NaOH to form alkaline picrate, stored in a dark amber reagent bottle. Alkaline picrate (1 mL) was added to 1 mL of diluted urine (1/10 v/v in deionized water). Optical density was measured at 500 nm for 30 min using a Shimadzu mini 1240 spectrophotometer. Creatinine levels also play a crucial role in assessing the suitability of urine samples for health risk assessment. The WHO recommends excluding samples with excessively low (<30 mg/dL Cr) or high (>300 mg/dL Cr) creatinine concentrations (35). Consequently, 62 urine samples were excluded from subsequent risk assessment studies due to these criteria.

### 2.7 Estimated dietary exposures

The probable daily intake of each mycotoxin was estimated from the measured urinary biomarker (34,40) using Equation (2).

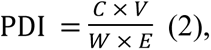

where C is the urinary biomarker concentration (µg/L), V is the mean volume of urine (1.5 L/day) produced daily by adults, W is body weight (kg), and E is the mean urinary excretion rate. The urinary excretion rate for FB1 is 0.5% (41) and 9.4 % for total ZEN (42).

Ethical clearance for the biomonitoring exercise was obtained from the Committee on Human Research and Publication Ethics (CHRPE), KNUST. All processes and activities were conducted according to the CHRPE guidelines and recommendations registered under CHRPE/AP/245/21 given for this study.

## 3.0 Results

### 3.1 Urine creatinine levels across the four regions

Analysis of creatinine levels in participants’ urine across the four regions revealed significant inter-regional variability (Table 1). The regional distribution of valid urine samples from study participants, collected for further analysis, is shown in Figure 1Sa. Figure 1Sb illustrates the percentage distribution of study participants across the study areas. Studies (43,44), have shown that spot urine creatinine estimations are equivalent to 24 h sample assessments. Generally, urine creatinine concentrations per 24 h for samples collected from females range between 6.79 and 19.09 mg/dL (45). With 120 samples, the Ashanti Region demonstrated a relatively narrow range of creatinine concentrations (4.76-91.40 mg/dL), suggesting a more homogenous population. The mean concentration of 85.61 mg/dL and 72.5 mg/dL standard deviation indicated moderate intra-regional variability. In contrast, the Greater Accra Region, with 67 samples, exhibited the highest mean creatinine concentration (145.97 mg/dL) and the widest range (11.34-465.43 mg/dL), indicative of more significant heterogeneity within the sampled population. The high standard deviation of 101.4 mg/dL further emphasized the considerable dispersion of creatinine levels within this region.

**Table 1.**
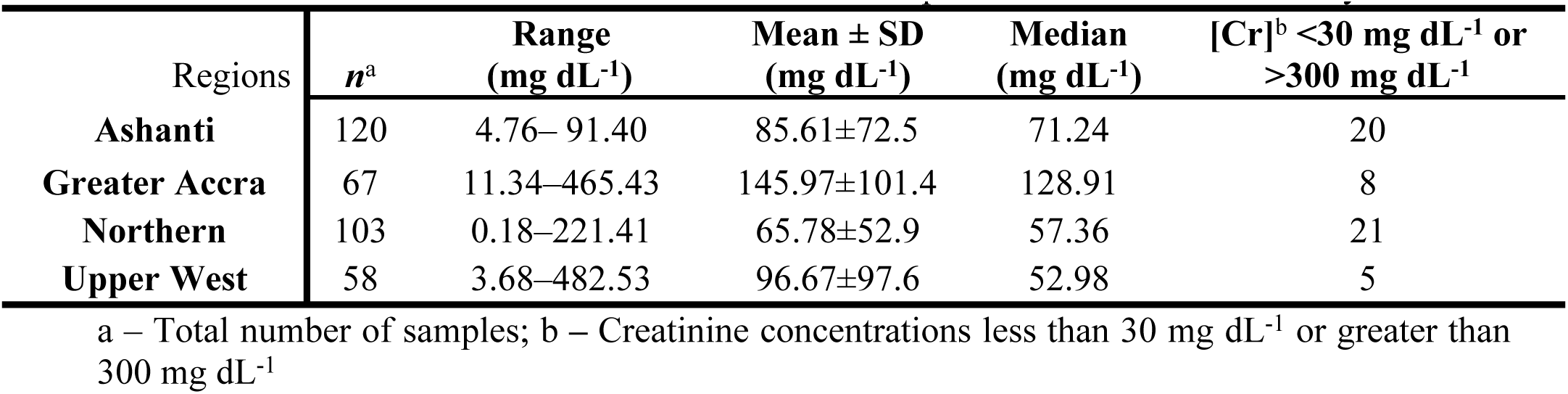
Creatinine concentrations in the human urine bio-samples collected across the study areas.

| Regions | *n*<sup>a</sup> | Range<br>(mg dL<sup>-1</sup>) | Mean ± SD<br>(mg dL<sup>-1</sup>) | Median<br>(mg dL<sup>-1</sup>) | [Cr]<sup>b</sup> <30 mg dL<sup>-1</sup> or<br>>300 mg dL<sup>-1</sup> |
| --- | --- | --- | --- | --- | --- |
| **Ashanti** | 120 | 4.76–91.40 | 85.61±72.5 | 71.24 | 20 |
| **Greater Accra** | 67 | 11.34–465.43 | 145.97±101.4 | 128.91 | 8 |
| **Northern** | 103 | 0.18–221.41 | 65.78±52.9 | 57.36 | 21 |
| **Upper West** | 58 | 3.68–482.53 | 96.67±97.6 | 52.98 | 5 |

a – Total number of samples; b – Creatinine concentrations less than 30 mg dL<sup>-1</sup> or greater than 300 mg dL<sup>-1</sup>

The Northern Region, represented by 103 samples, displayed a mean creatinine concentration of 65.78 mg/dL with a standard deviation of 52.9 mg/dL. A significant number of 21 samples exhibited values outside the World Health Organization (WHO) recommended range (less than 30 mg/dL or greater than 300 mg/dL) for exposure assessment. This suggests the potential presence of underlying health conditions or confounding factors influencing renal function in a subset of this population. With the smallest sample size (58), the Upper West Region demonstrated the highest maximum concentration (482.53 mg/dL) and the most significant variability among the regions. The mean concentration of 96.67 mg/dL and a standard deviation of 97.6 mg/dL underscored significant heterogeneity in this population, likely influenced by many factors

### 3.2 Age-dependent patterns of mycotoxin biomarker levels in human female urine

Table 2 presented a comprehensive overview of mycotoxin biomarker concentrations in human female urine samples across different age groups. Fumonisin B1 (FB1) median levels exhibited minor variations across age groups, with the 55+ group showing the highest value at 89.65 ng/mL. Creatinine-adjusted levels demonstrated an apparent age-dependent increase, culminating in the highest values in the 55+ group (174.53 ng/g Cr). Fumonisin B2 (FB2) median levels exhibited a significant age-related increase, peaking in the 55+ group (1117.80 ng/mL). This trend mirrored the creatinine-adjusted levels, reaching their highest values in the 55+ group (1398.6 ng/g Cr). Zearalenone (ZEN) median levels were highest in the 55+ age group (69.86 ng/mL). Creatinine-adjusted levels also demonstrated the highest values in this age group (113.33 ng/g Cr). α-Zearalenol (α-ZEL) median levels were highest in the 20-54 age group (14.60 ng/mL). However, creatinine-adjusted levels showed a different trend, peaking in the 10-19 age group (37.77 ng/g Cr). β-Zearalenol (β-ZEL) levels mirrored α-ZEL, with the highest median in the 55+ group (28.35 ng/mL). However, like α-ZEL, creatinine-adjusted levels peaked in the 10-19 age group (31.45 ng/g Cr). Zearalenone 14-Glucuronide (ZEN-14-GlcA) median levels remained relatively consistent across age groups, with the highest level observed in the 10-19 age group (3.80 ng/mL). Creatinine-adjusted levels reached their highest point in the 20-54 age group (9.38 ng/g Cr).

**Table 2.** Occurrence of mycotoxin biomarker concentrations in human female urine samples collected from the study areas.

| Age Groups | Central tendencies of biomarker concentrations |  |  |  |  |  |  |
| --- | --- | --- | --- | --- | --- | --- | --- |
|  |  |  |  | Urinary biomarker levels |  | Creatinine-adjusted urinary biomarker levels |  |
|  | *n*<sup>a</sup> | N%<sup>b</sup> | <LOD<sup>c</sup> | Range<br>(ng/mL) | Median<br>(ng/mL) | Range<br>(ng/g Cr) | Median<br>(ng/g Cr) |
|  | Fumonisin B1 (FB1) |  |  |  |  |  |  |
| 10-19 | 29 | 37.9% | 18 | 49.48–115.81 | 81.99 | 40.37–350.60 | 84.76 |
| 20-54 | 236 | 37.7% | 147 | 3.51–1302.8 | 78.80 | 9.47–9778.9 | 104.11 |
| 55+ | 29 | 44.8% | 16 | 1.88–209.99 | 89.65 | 1.55–1794.8 | 174.53 |
| Fumonisin B2 (FB2) |  |  |  |  |  |  |  |
| 10-19 | 29 | 24.1% | 22 | 2.66–17844.7 | 164.57 | 5.18–7755.8 | 171.30 |
| 20-54 | 236 | 27.9% | 170 | 0.10–16365 | 116.50 | 0.29–35971 | 125.3 |
| 55+ | 29 | 41.4% | 18 | 0.41–7217.5 | 1117.80 | 0.19–48188.9 | 1398.6 |
| Zearalenone (ZEN) |  |  |  |  |  |  |  |
| 10-19 | 29 | 6.9% | 27 | 14.24–45.02 | 26.63 | 19.57–32.45 | 26.01 |
| 20-54 | 236 | 15.3% | 200 | 1.89–134.14 | 16.47 | 2.24–273.25 | 22.39 |
| 55+ | 29 | 17.2% | 24 | 21.1–222.22 | 69.86 | 0.36–846.23 | 113.33 |
| α- Zearalenol (α-ZEL) | | | | | | | |
| 10-19 | 29 | 44.8% | 16 | 8.72–30.42 | 13.66 | 8.32–280.29 | 37.77 |
| 20-54 | 236 | 45.3% | 129 | 7.39–97.65 | 14.60 | 3.94–812.56 | 22.39 |
| 55+ | 29 | 48.3% | 15 | 7.14–35.56 | 16.49 | 0.31–295.35 | 15.73 |
| β- Zearalenol (β-ZEL) | | | | | | | |
| 10-19 | 29 | 65.5% | 10 | 4.63–175.54 | 19.11 | 6.67–265.89 | 31.45 |
| 20-54 | 236 | 60.2% | 94 | 2.50–109.25 | 16.69 | 1.54–1750.74 | 26.81 |
| 55+ | 29 | 51.7% | 14 | 7.16–88.36 | 28.35 | 0.73–940.04 | 25.93 |
| Zearalenone 14-Glucuronide (ZEN-14-GlcA) |  |  |  |  |  |  |  |
| 10-19 | 29 | 13.8% | 25 | 3.79–11.35 | 3.80 | 1.65– 29.66 | 5.78 |
| 20-54 | 236 | 15.3% | 200 | 3.76–15.93 | 3.89 | 1.97–116.55 | 9.38 |
| 55+ | 29 | 17.2% | 24 | 3.78–11.40 | 3.81 | 3.54–40.49 | 4.11 |

a -Sample size; b- percentage of positive samples; c- limit of detection

### 3.3 Statistical distributions and central tendencies of FumB1 and Zearalenone biomarkers

Table 3 outlines biomarkers’ statistical distributions and central tendencies for fumonisin B1 (FB1) and Zearalenone (ZEN) exposure. Detailed descriptions of the distributions are presented in the Supplementary material (Supplement S2). FB1 concentrations are characterized by a mode of 65.8 µg/L and a median of 76.9 µg/L. The 5^th^ and 95^th^ percentiles were 25.2 µg/L and 181 µg/L, respectively. ZEN concentrations recorded a mode of 0.01 µg/L and a median value of 19.8 µg/L. The 5^th^ percentile was 0.82 µg/L, while the 95th percentile was 87.7 µg/L. α-zearalenol (α-ZEL), also recorded a mode of 12.9 µg/L and a median value of 15.1 µg/L with 5^th^ and 95^th^ percentile concentrations being 9.05 and 32.3 µg/L respectively. The 5^th^ percentile β-zearalenol (β-ZEL) concentration was 5.12 µg/L, the 95^th^ was 83.9 µg/L and the mode and median concentrations were 8.12 and 18.6 µg/L respectively. The modal ZEN-14-GlcA value was similar to ZEN, while the 5th and 9th percentile concentrations were 4.81 µg/L and 14.5 µg/L, respectively. Total ZEN biomarkers (tZENBM) recorded a mode of 11.7 µg/L and a median value of 28.7 µg/L, while its 5^th^ percentile concentration was 4.50 µg/L urine.

**Table 3:**
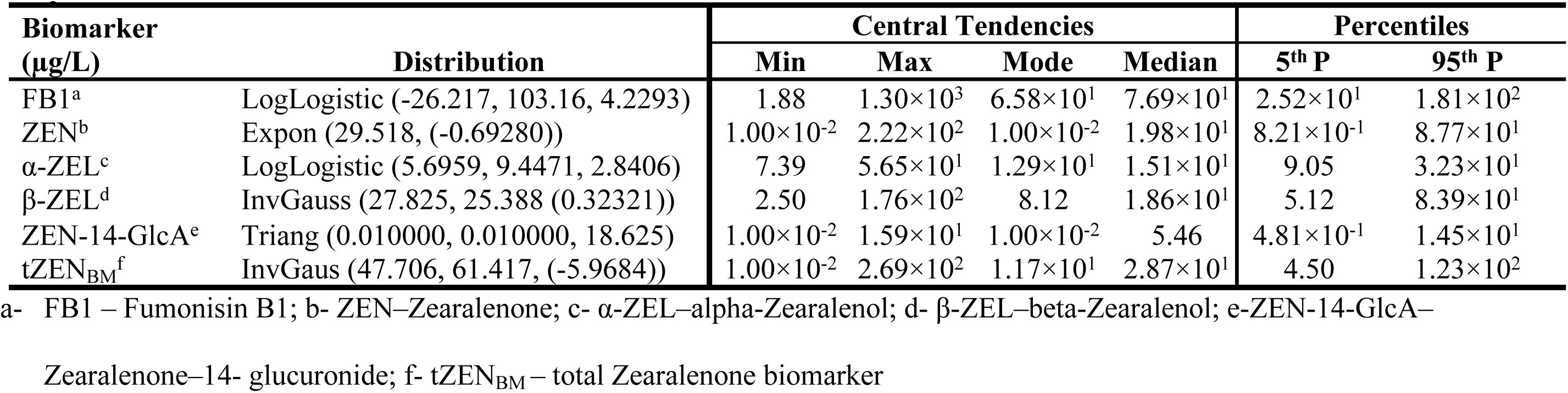
Statistical distributions and central tendencies of the concentrations of biomarkers of fumonisin B1 and Zearalenone exposure.

### 3.4 Age-specific differences in mycotoxin biomarker distribution

Table 4 presented the distribution of Fumonisin B1 (FB1) and Zearalenone (ZEN) biomarkers across three age groups: 10-19, 20-54, and 55+.

**Table 4:** Elements of exposure of Fumonisin B1 (FB1), zearalenone and its metabolite biomarkers (tZEN), statistical distributions and central tendency metrics.

| Element | Distribution | Central Tendencies |  |  |  | Percentiles |  |
| --- | --- | --- | --- | --- | --- | --- | --- |
|  |  | Min | Max | Mode | Median | 5<sup>th</sup> | 95<sup>th</sup> |
| 10-19 age group |  |  |  |  |  |  |  |
| FB1<sup>a</sup> (µg/L) | Uniform (42.121, 123.182) | 4.94×10<sup>1</sup> | 1.16×10<sup>2</sup> | 7.44×10<sup>1</sup> | 8.27×10<sup>1</sup> | 4.62×10<sup>1</sup> | 1.19×10<sup>2</sup> |
| ZEN<sup>b</sup> (µg/L) | Pareto (0.44662, 0.010000) | 1.00×10<sup>-2</sup> | 4.50×10<sup>1</sup> | 1.00×10<sup>-2</sup> | 4.27×10<sup>-1</sup> | 1.00×10<sup>-2</sup> | 8.18 |
| α-ZEL<sup>c</sup> (µg/L) | Uniform (6.5479, 32.594) | 8.72 | 3.04×10<sup>1</sup> | 8.97 | 1.96×10<sup>1</sup> | 7.85 | 3.19×10<sup>1</sup> |
| β-ZEL<sup>d</sup> (µg/L) | Expon 45.155, (2.7735)) | 5.47 | 1.76×10<sup>2</sup> | 2.77 | 3.27×10<sup>1</sup> | 4.99 | 1.32×10<sup>2</sup> |
| ZEN-14-GlcA<sup>e</sup> (µg/L) | Pareto (0.37008, 0.010000) | 1.00×10<sup>-2</sup> | 1.14×10<sup>2</sup> | 1.00×10<sup>-2</sup> | 6.51×10<sup>-2</sup> | 1.15×10<sup>-2</sup> | 3.28×10<sup>1</sup> |
| tZEN<sub>BM</sub><sup>f</sup> (µg/L) | Expon (50.182, (-2.3796)) | 1.00×10<sup>-2</sup> | 2.01×10<sup>2</sup> | 1.49×10<sup>1</sup> | 3.24×10<sup>1</sup> | 1.94×10<sup>-1</sup> | 1.47×10<sup>2</sup> |
| Bw<sup>g</sup> (kg) | Laplace (55,8.1748) | 41 | 80.4 | 55 | 55 | 41.7 | 68.3 |
| 20-54 age group |  |  |  |  |  |  |  |
| FB1 (µg/L) | LogLogistic (-16.044, 91.740, 3.5344) | 3.51 | 1.30×10<sup>3</sup> | 6.18×10<sup>1</sup> | 7.57×10<sup>1</sup> | 2.38×10<sup>1</sup> | 1.95×10<sup>2</sup> |
| ZEN (µg/L) | Expon (26.494, (0.98369)) | 1.90 | 1.34×10<sup>2</sup> | 9.84×10<sup>-1</sup> | 1.94×10<sup>1</sup> | 2.34 | 8.04×10<sup>1</sup> |
| α-ZEL (µg/L) | LogLogistic (5.5920, 9.5478, 2.9475) | 7.39 | 5.65×10<sup>1</sup> | 1.31×10<sup>1</sup> | 1.51×10<sup>1</sup> | 9.11 | 3.15×10<sup>1</sup> |
| β-ZEL (µg/L) | Kumaraswamy (0.69723, 2.2564, 109.53) | 2.50 | 1.02×10<sup>2</sup> | 2.50 | 1.84×10<sup>1</sup> | 2.97 | 7.13×10<sup>1</sup> |
| ZEN-14-GlcA (µg/L) | Lognorm (60.610, 8756.2, 3.7787) | 3.78 | 1.59×10<sup>1</sup> | 3.78 | 4.20 | 3.78 | 7.88×10<sup>1</sup> |
| tZEN<sub>BM</sub> (µg/L) | Kumaraswamy (0.69928, 2.1616, 143.33) | 2.16 | 1.36×10<sup>2</sup> | 2.16 | 1.84×10<sup>1</sup> | 3.01 | 1.06×10<sup>2</sup> |
| Bw (kg) | Extvalue (59.971,11.089) | 41.0 | 116.5 | 59.9 | 64.0 | 47.8 | 92.9 |
| 55+ age group |  |  |  |  |  |  |  |
| FB1 (µg/L) | Extvalue (91.1733,23.2475) | 1.88 | 1.12×10<sup>2</sup> | 9.17×10<sup>1</sup> | 8.26×10<sup>1</sup> | 2.21×10<sup>1</sup> | 1.17×10<sup>2</sup> |
| ZEN (µg/L) | Pareto (0.17186, 0.010000) | 1.00×10<sup>-2</sup> | 2.22×10<sup>2</sup> | 1.00×10<sup>-2</sup> | 5.64×10<sup>-1</sup> | 1.35×10<sup>-2</sup> | 3.72×10<sup>4</sup> |
| α-ZEL (µg/L) | Extvalue (13.8189, 3.6319) | 9.59 | 2.56×10<sup>1</sup> | 1.38×10<sup>1</sup> | 1.52×10<sup>1</sup> | 9.83 | 2.46×10<sup>1</sup> |
| β-ZEL (µg/L) | Expon (19.619, (5.6484)) | 7.156 | 7.03×10<sup>1</sup> | 5.65 | 1.92×10<sup>1</sup> | 6.66 | 6.42×10<sup>1</sup> |
| ZEN-14-GlcA (µg/L) | Pareto (0.23121, 0.010000) | 1.00×10<sup>-2</sup> | 1.14×10<sup>1</sup> | 1.00×10<sup>-2</sup> | 2.00×10<sup>-1</sup> | 1.25×10<sup>-2</sup> | 4.24×10<sup>3</sup> |
| tZEN<sub>BM</sub> (µg/L) | Expon (53.548, (-3.1399)) | 1.00×10<sup>-2</sup> | 2.69×10<sup>2</sup> | 1.74×10<sup>1</sup> | 3.39×10<sup>1</sup> | 1.00×10<sup>-2</sup> | 1.57×10<sup>2</sup> |
| Bw (kg) | Uniform (35.935,87.565) | 38 | 85.5 | 69.7 | 61.8 | 38.5 | 84.9 |

a- FB1 – Fumonisin B1; b- ZEN – Zearalenone; c- α-ZEL – alpha-Zearalenol; d- β-ZEL – beta-Zearalenol; e- ZEN-14-GlcA – Zearalenone – 14- glucuronide; f- tZEN<sub>BM</sub> – total Zearalenone biomarkers; g- Bw – Body weight

#### Age Group 10-19

In this age group, FB1 exposure exhibited a uniform distribution with a high median of 74.4 ng/mL, suggesting consistent exposure across the population. However, significant variability was observed, with the 5^th^ and 95^th^ percentiles ranging from 46.2 ng/mL to 119 ng/mL. Zearalenone (ZEN) exposure followed a Pareto distribution, indicating that a small proportion of individuals experienced substantially higher exposure levels (maximum 45 ng/mL) compared to the majority (median 0.43 ng/mL). α-ZEL demonstrated a uniform distribution with a median of 19.6 ng/mL, while β-ZEL exhibited an exponential distribution with a median of 32.7 ng/mL. ZEN-14-Glucose (ZEN-14-GlcA) also followed a Pareto distribution, mirroring the trend observed for ZEN, with a median of 0.065 ng/mL. Total ZEN biomarkers (tZEN_BM_) exhibited an exponential distribution, with a median value of 32.4 ng/mL, suggesting higher concentrations in a smaller population subset. Body weight (BW) in this group followed a Laplace distribution, with a peak around the median value of 55 kg, indicating that most individuals’ body weights were clustered around this value.

#### Age Group 20-54

The FB1 exposure followed a log-logistic distribution for this age bracket, suggesting higher concentrations among a smaller population subset. The 5^th^ and 95^th^ percentiles further supported the observation, ranging from 23.8 ng/mL to 195 ng/mL. ZEN exposure in this group exhibited an exponential distribution, with a median of 75.7 ng/mL and a maximum value of 1, 300 ng/mL, indicating that while most individuals had lower exposure levels, some experienced significantly higher levels. α-ZEL demonstrated a log-logistic distribution with a median of 15.1 ng/mL. β-ZEL followed a Kumaraswamy distribution, highlighting higher concentration levels among fewer individuals and reflecting substantial variability in exposure across the group, with 18.4 ng/mL at the median. ZEN-14-Glucose (ZEN-14-GlcA) displayed a lognormal distribution, suggesting that while the majority had lower exposure levels, a small proportion experienced considerably higher levels, with a median value of 4.20 ng/mL. The total ZEN biomarkers (tZEN_BM_) also followed a Kumaraswamy distribution, indicating a similar trend to β-ZEL, with a value of 18.4 ng/mL at the median.

#### Age Group 55+

Fumonisin B1 (FB1) exposure followed a General Extreme Value distribution in this older age group, indicating a central clustering of exposure levels with moderate variation (82.6 ng/mL). Zearalenone (ZEN) exposure exhibited a normal distribution, suggesting a symmetrical distribution around the mean exposure level of 0.56 ng/mL. α-ZEL also showed an extreme value distribution, and a median of 15.2 ng/mL, suggesting a central clustering and significantly higher exposure levels. β-ZEL followed an exponential distribution, higher exposure levels concentrated among a small subset of the population, with a median value of 19.2 ng/mL. ZEN-14-Glucuronide (ZEN-14-GlcA) recorded a median value of 0.20 ng/mL, and also followed a Pareto distribution, indicating that exposures were prevalent in fewer individuals. Total ZEN biomarkers (tZEN_BM_) exhibited an exponential distribution, similar to β-ZEL, indicating central clustering with significant spread, with a median value of 33.9 ng/mL. Body weight (BW) in this group reflected a uniform distribution, indicating that body weights were symmetrically distributed around a median value of 61.8 kg.

The study, therefore, highlighted the variability and the potential health implications of exposure to these mycotoxins across different age groups. FB1 in the 10-19 age group demonstrated relatively consistent exposure, while ZEN and its metabolites in all age groups exhibited significant variability and skewed distributions. The observation suggests that a smaller proportion of individuals within each age group may be experiencing substantially higher exposure levels.

### 3.5 Variations in mycotoxin exposure

For Fumonisin B1 (FB1), the data revealed no particular age-related trend in maximum probable daily intake (PDI). The 20-54 year group exhibited the highest maximum PDI value at 39.5 µg/kg(bw)-day, followed by the 10-9 year group with 38.3 µg/kg(bw)-day, and a declined for those aged 55+. At the 5^th^ percentile, the older age groups recorded similar PDI, higher than the younger demography but the differences in PDI were more pronounced at the 95^th^ percentile and median. At the median, estimated FB1 exposures were higher for the adolescents and teens while at the 95^th^ percentile, adults (20–54) and the elderly (55+) were more exposed. The trend for ZEN indicated an exceptionally high median PDI for the 20-54 age group, which was recorded at 4.17 and dropped markedly for other age groups. Similar to FB1, the 5^th^ and 95^th^ percentile values followed no discernible age-related trend, except that the 95^th^ percentile exposures were expectedly higher than 5^th^ exposures.

α-Zearalenol (α-ZEL) presented the highest PDI in the 10-19 year group at 378 µg/kg(bw)-day, decreasing as age increased. Although α-ZEL had relatively low mode and median values, these also showed slight reductions with age. The 5^th^ and 95^th^ percentile values were low and decreased subtly with age. Similarly, for β-Zearalenol (β-ZEL), the maximum PDI peaked in the 10-19 age group at 989 µg/kg(bw)-day, with a progressive decrease across older age groups. The mode and median values of β-ZEL similarly exhibited minor decreases with age, while the 5^th^ and 95^th^ percentiles showed slight reductions.

With Zearalenone-14-glucuronide (ZEN-14-GlAc), the maximum PDI were exceptionally high across the ages but more pronounced in the 55+ followed by the 10-19 year groups. The exceptionally high values could be attributed to possible outlier influence. While at the median, the 20-54 year group seems significantly more exposed, at the 95^th^ percentile, the elderly recorded higher PDI than the other two age groups. For Total ZEN biomarkers (t-ZEN), the maximum PDI was the highest for the 10-19 year group and least in the 20-54 year group. The median and 95^th^ percentile PDI underscored this trend, indicating low PDI (6.38 µg/kg(bw)-day) in the adult age group in contrast with the estimates recorded for the 10-19 and 55+ age groups respectively (Table 5).

**Table 5:** Estimated Probable Daily Intake (PDI) µg/kg bw of the dietary mycotoxins from urinary biomarker data.

| Age Groups<br>(years) | Central tendencies |  |  |  | Percentiles |  |
| --- | --- | --- | --- | --- | --- | --- |
|  | Min | Max | Mode | Median | 5<sup>th</sup> | 95<sup>th</sup> |
| **Fumonisin B1 (FB1)** |  |  |  |  |  |  |
| 10-19 | 0 | $3.83 \times 10^1$ | 1.03 | 1.13 | $6.11 \times 10^{-2}$ | 1.78 |
| 20-54 | 0 | $3.95 \times 10^1$ | $6.26 \times 10^{-1}$ | $8.72 \times 10^{-1}$ | $2.62 \times 10^{-1}$ | 2.37 |
| 55+ | 0 | 2.82 | $9.44 \times 10^{-1}$ | $9.79 \times 10^{-1}$ | $2.63 \times 10^{-1}$ | 1.84 |
| **Zearalenone (ZEN)** |  |  |  |  |  |  |
| 10-19 | 0 | $1.47 \times 10^{15}$ | $2.97 \times 10^{-2}$ | $1.23 \times 10^{-2}$ | $2.84 \times 10^{-2}$ | 2.15 |
| 20-54 | $9.39 \times 10^{-2}$ | $1.12 \times 10^2$ | $3.53 \times 10^{-1}$ | 4.17 | $5.00 \times 10^{-1}$ | $1.81 \times 10^1$ |
| 55+ | $1.62 \times 10^{-3}$ | $4.34 \times 10^{30}$ | $3.02 \times 10^{-3}$ | $1.34 \times 10^{-1}$ | $3.13 \times 10^{-3}$ | $8.88 \times 10^4$ |
| **α-Zearalenol (α-ZEL)** |  |  |  |  |  |  |
| 10-19 | 0 | $3.78 \times 10^2$ | 5.33 | 5.02 | 1.98 | 8.65 |
| 20-54 | $7.35 \times 10^{-1}$ | $1.01 \times 10^2$ | 2.67 | 3.33 | 1.77 | 7.30 |
| 55+ | $8.84 \times 10^{-1}$ | $1.87 \times 10^1$ | 2.93 | 3.55 | 1.99 | 6.81 |
| **β-Zearalenol (β-ZEL)** |  |  |  |  |  |  |
| 10-19 | 0 | $9.89 \times 10^2$ | 1.46 | 8.46 | 1.28 | $3.51 \times 10^1$ |
| 20-54 | $1.97 \times 10^{-1}$ | $3.41 \times 10^1$ | $7.30 \times 10^{-1}$ | 3.97 | $6.45 \times 10^{-1}$ | $1.61 \times 10^1$ |
| 55+ | $9.15 \times 10^{-1}$ | $7.43 \times 10^1$ | 2.07 | 4.53 | 1.45 | $1.64 \times 10^1$ |
| **Zearalenone – 14- glucuronide (ZEN-14- GlAc)** |  |  |  |  |  |  |
| 10-19 | 0 | $1.52 \times 10^{12}$ | $2.77 \times 10^{-3}$ | $1.70 \times 10^{-2}$ | $2.92 \times 10^{-3}$ | 8.49 |
| 20-54 | $2.70 \times 10^{-1}$ | $8.60 \times 10^4$ | $8.52 \times 10^{-1}$ | 1.01 | $6.35 \times 10^{-1}$ | $1.74 \times 10^1$ |
| 55+ | $1.62 \times 10^{-3}$ | $2.08 \times 10^{20}$ | $2.92 \times 10^{-3}$ | $4.77 \times 10^{-2}$ | $2.84 \times 10^{-3}$ | $1.01 \times 10^3$ |
| **Total ZEN biomarkers (t-ZEN)** |  |  |  |  |  |  |
| 10-19 | 0 | $3.32 \times 10^2$ | $2.86 \times 10^{-1}$ | 8.37 | $4.93 \times 10^{-2}$ | $3.92 \times 10^2$ |
| 20-54 | $2.23 \times 10^{-1}$ | $4.97 \times 10^1$ | $6.31 \times 10^{-1}$ | 6.38 | $4.16 \times 10^{-1}$ | $2.42 \times 10^1$ |
| 55+ | 0 | $1.85 \times 10^2$ | 0 | 7.89 | 0 | $3.94 \times 10^1$ |

The maximum PDI values for all mycotoxins generally decreased across age groups. Both mode and median values showed lowering PDI for those aged 20-54, although the reductions were less pronounced than the maximum values. The percentile values, especially the 95^th^ percentile, significantly highlighted the fluctuating PDI with age, indicating a higher variation in exposure levels among the sampled population.

## 4.0 Discussion

### 4.1 Creatinine concentrations and kidney function

The current data (Table 1) indicated that while the Ashanti Region showed a narrow range of creatinine concentrations (4.76-91.40 mg/dL), the Greater Accra Region exhibited the highest mean (145.97 mg/dL) and the widest range (11.34-465.43 mg/dL) creatinine concentration.

Recent research on regional variations in creatinine concentrations and kidney function has highlighted similar trends to the observations in different parts of Ghana. According to a study published by the Global Burden of Disease (GBD), there are significant regional disparities in chronic kidney disease (CKD) prevalence and creatinine levels worldwide (46). Thus, the high variability in creatinine levels within regions is often associated with population heterogeneity and underlying health conditions. Comparatively, the GBD study found that regions with higher mean creatinine levels and more significant variability often had diverse populations and contributing factors such as socioeconomic status, access to healthcare, and environmental influences (47). The range of creatinine concentrations determined in this study across the four study areas exceeded the clinically suggested range (6.79 -19.09 mg/dL) suggesting underlying problems with renal function, particularly in the Greater Accra and Upper West Regions.

### 4.2 Mycotoxin concentrations as biomarkers in human samples

Data from Table 2 observed that age-dependent variations in mycotoxin levels were the highest in the 55+ group. Indeed, this observation is consistent with the literature (48) suggesting bioaccumulation of mycotoxins with age. For Fumonisin B1 (FB1), higher levels in older individuals are consistent with findings of increased accumulation with age. Similarly, the significant age-related increase in FB1 levels supports the notion of age-related metabolic changes impacting mycotoxin absorption and excretion. Zearalenone (ZEN) and its metabolites, including α-Zearalenol and β-Zearalenol, frequently exhibit higher concentrations in older populations, corroborating the trend of highest median levels in the 55+ group. While Zearalenone 14-Glucuronide (ZEN-14-GlcA) levels appear relatively consistent across age groups, the observed peaks in specific age groups, such as the 20-54 group, suggest that specific mycotoxin metabolite levels can vary significantly with age, as supported by other research findings (48). The findings obtained from this current study favorably compare with standard methodologies in mycotoxin research, emphasizing the importance of urinary biomarkers for evaluating human exposure (47). FB1 emerged as the most abundant mycotoxin to frequently occur as an element of exposure in the study (Table 4) peaking in the 55+ age group (91.7 ng/mL). The estimated mycotoxin PDI from urinary biomarker analysis however did not emphasize this trend, indicating that intake and subsequent exposures can be attributed to several factors. While prolonged dietary exposure may play a significant role in exposures, the mass of food consumed significantly influences exposure levels as well. Also, age-related metabolic changes can lead to higher body retention and accumulation of mycotoxins (49) Environmental factors, such as inadequate storage conditions, including high humidity and temperature, can increase mycotoxin contamination in food (50). Age-related physiological changes, such as compromised liver function in older individuals, can impair the body’s ability to metabolize and excrete mycotoxins (51).

### 4.3 Patterns of Diverse Mycotoxin Distributions

Table 3 presented key distributions illustrating the spread of mycotoxins within the study communities. Zearalenone 14-Glucuronide (ZEN-14-GlcA) followed a Triangular Distribution, establishing limited sample data is available (52). Such distributions fit a range defined by a minimum, maximum, and mode, providing a practical approximation when exact distributions are difficult to ascertain. β-Zearalenol (β-ZEL) and total ZEN biomarkers (tZEN_BM_) typically followed an Inverse Gaussian Distribution, characterized by a heavy tail and a peak at lower concentrations, a typical pattern in life and health sciences (53). Zearalenone (ZEN) also followed Exponential Distribution, reflecting the dynamics of short-lived mycotoxins in human samples (54). Fumonisin B1 (FB1) and α-Zearalenol (α-ZEL) exhibited a log-logistic distribution, which is particularly well-suited for describing the skewed concentration levels of fumonisins in biological samples. The observation aligns with studies confirming that FB1 contamination in food products leads to skewed concentration distribution in human samples. Furthermore, the binding behaviour of ZEN and its metabolites (including α-ZEL) with serum albumins highlights the variability in concentration levels across species, supporting the observed patterned distribution in biological samples (55–57).

### 4.4 Demographic attributes of mycotoxin exposure and implications for public health

Table 4 underscored a critical point of effectively addressing mycotoxin exposure necessitated a nuanced consideration of how demographic classifications influenced individual experiences. Such a conscious application of demographic factors within exposure pathways would have significantly impacted health outcomes and the effectiveness of interventions.

#### Age Group 10-19

This age group frequently exhibited higher mycotoxin exposure levels, particularly for zearalenone (ZEN), with a notable correlation observed between higher ZEN exposure and lower body weights (58). Younger individuals with lower body weights often exhibit higher exposure to mycotoxins, such as ZEN, as with other hazards (59) as indicated by Equation (2). The relationship between exposure and effect may be attributed to various factors, including dietary differences and metabolic rates (60). Significant variability in ZEN exposure levels was observed within this age group, indicating that some individuals were at substantially higher risk than others (61).

#### Age Group 20-54

This age group exhibited a wide range and variability of mycotoxin exposures, likely influenced by lifestyle and dietary habits, with some individuals displaying higher concentrations of ZEN and fumonisins (62). ZEN exposure often followed an exponential distribution, with a smaller subset experiencing significantly higher mycotoxin levels (63). Adults within this group exhibited a broad spectrum of body weights and mycotoxin exposure levels (64). Thus, the study suggested an exponential distribution of mycotoxin exposure, with specific subsets showing significantly higher concentrations. This group demonstrated a peak in body weight of around 67.1 kg, with considerable variations potentially reflecting diverse lifestyles and dietary habits (62).

#### Older Adults (55+)

Older adults exhibited more uniform exposure patterns, often clustering around median values for ZEN and fumonisins, suggesting consistent exposure over time (65). The body weights in older adults were symmetrically distributed around 61.8 kg, suggesting to improved dietary awareness and weight management (66).

Across all age groups, a notable correlation existed between lower body weights and higher exposure to mycotoxins, particularly ZEN. This correlation was especially significant in younger individuals (10-19 y), highlighting the need for targeted public health interventions(67).

### 4.5 Mycotoxin exposure of fumonisin B1 and zearalenone metabolites in varied age demographics

Table 5 reveals a significant decrease in maximum probable daily intake (PDI) and consistent exposure values for Fumonisin B1 (FB1) with increasing age. Specifically, the maximum PDI decreased from 38.3 µg/kg(bw)-day in the 10-19 age group to 2.82 µg/kg(bw)-day in the 55+ age group. The data aligns with research demonstrating lower dietary fumonisin intake in older populations due to diversified and less frequent consumption of potentially contaminated foods (56). The trend differs for ZEN, with high exposure in the 20-54 age group (median PDI of 4.17 µg/kg(bw)-day) peaking in the 55+ age group at the 95^th^ percentile (68). This trend repeated with all ZEN metabolites, including α-ZEL, which shows a decrease from a maximum PDI of 378 µg/kg(bw)-day in the 10-19 age group, and β-ZEL reducing from 989 µg/kg(bw)-day, across older age groups. t-ZEN also had a maximum PDI of 332 µg/kg(bw)-day for the 10-19 age group and declined accordingly. Although outliers likely influenced ZEN-14-GlcA maximum levels, PDI estimates at other central tendencies followed the general reduction trend with age. In all these cases, younger age demographics face more significant exposure risks due to their higher frequency of cereal consumption. At the same time, adult populations exhibit reduced mycotoxin intake overall, likely due to increased awareness of dietary variety and adherence to regulatory measures(69,70).

## 5.0 Limitations of the study

One significant limitation of this study is the reliance on single-spot urine samples per participant. While this approach is practical for assessing population-level exposure trends, it may not fully capture the variability in individual exposures over time. Furthermore, the study’s exclusive focus on females within the cereal value chain necessitates caution when generalizing findings to populations with diverse gender compositions. The reason is that males and females often exhibit distinct dietary patterns, which can significantly influence their exposure to dietary hazards.

Another limitation lies in the probabilistic estimation of probable daily intake, which relied on assumptions regarding excretion ratios without accounting for individual variations in metabolism and mycotoxin excretion. The exclusion of invalid urine samples further reduced the sample size, potentially impacting the study’s statistical power.

As a remedy, median values were reported from probabilistic estimation approaches. While a more frequent sampling regimen could provide deeper insights into the variability of chronic exposures, the study’s primary objective of establishing population exposure levels within the cereal value chain was adequately addressed through the chosen sampling method.

## 6.0 Conclusion

The biomonitoring study revealed significant regional variations in urinary biomarker occurrence and levels in females in the cereal value chain. Each biomarker followed specific distribution patterns such as Exponential, Log-logistic and Gaussian distributions, resulting in unique age-related probable daily intake for FB1 and ZEN and its metabolites. Creatinine-adjusted biomarker levels, meant to offer insight into renal function, were consistent with the observed regional mycotoxin exposures. The results suggest a homogeneity in creatinine clearance among the study participants in the Ashanti region, a heterogeneity for Greater Accra and deviations pointing to serious underlying conditions for the other two regions. Urinary biomarker levels, particularly FB1, are high, increasing for older age groups in patterns consistent with age-related accumulation. ZEN and its metabolites also presented age-dependent variations in their occurrence and concentrations, and younger individuals presented higher exposures. The study reveals the importance of age-related, demography and regional data considerations for conducting robust mycotoxin exposure risk assessment. The outcome of this biomonitoring exercise highlights the need for targeted public health interventions to mitigate the risks due to mycotoxin exposures in different populations.

### Data availability

Supplementary data related to this study can be accessed online at https://doi.org/10.17632/v6n8r3zy2f.3

## Data Availability

All the relevant data are within the manuscript and its Supporting Information files. A URL is also provided to easily access the Supporting Information files.

## Notes

### Competing Interest Statement

The authors have declared no competing interest.

### Funding Statement

The author(s) received no specific funding for this work.

### Author Declarations

Ethical clearance for this study was sought from the Committee on Human Research and Publication Ethics (CHRPE), Kwame Nkrumah University of Science and Technology (KNUST), Kumasi, Ghana. The clearance is registered under CHRPE/AP/245/21. A copy of the ethical clearance has been attached.

## Reference

1. Fan K, Xu J, Jiang K, Liu X, Meng J, Di Mavungu, JD, et al. Determination of multiple mycotoxins in paired plasma and urine samples to assess human exposure in Nanjing, China. Environ … [Internet]. 2019;248((2019)):865–73. Available from: https://www.sciencedirect.com/science/article/pii/S0269749118353089

2. Huang Q, Jiang K, Tang Z, Fan K, Meng J, Nie D, et al. Exposure Assessment of Multiple Mycotoxins and Cumulative Health Risk Assessment: A Biomonitoring-Based Study in the Yangtze River Delta, China. Toxins (Basel). 2021;13(2):1–15.

3. Arce-López B, Lizarraga E, Vettorazzi A, González-Peñas E. Human biomonitoring of mycotoxins in blood, plasma and serum in recent years: A review. Toxins (Basel). 2020;12(3):1–32.

4. Ali N, Degen GH. Urinary biomarkers of exposure to the mycoestrogen zearalenone and its modified forms in German adults. Arch Toxicol [Internet]. 2018;92(8):2691–700. Available from: 10.1007/s00204-018-2261-5

5. Torres O, Matute J, Gelineau-Van Waes J, Maddox JR, Gregory SG, Ashley-Koch AE, et al. Human health implications from co-exposure to aflatoxins and fumonisins in maize-based foods in Latin America: Guatemala as a case study. World Mycotoxin J. 2015;8(2):143–59.

6. Föllmann W, Ali N, Blaszkewicz M, Degen GH. Biomonitoring of Mycotoxins in Urine: Pilot Study in Mill Workers. J Toxicol Environ Heal - Part A Curr Issues [Internet]. 2016;79(22–23):1015–25. Available from: 10.1080/15287394.2016.1219540

7. Chen C, Mitchell NJ, Gratz J, Houpt ER, Gong Y, Egner PA, et al. Exposure to aflatoxin and fumonisin in children at risk for growth impairment in rural Tanzania. Environ Int. 2018;115:29–37.

8. IARC IA for R on C. Agents Classified by the IARC Monographs, Volumes 1 – 120. Vol. 7, IARC Monographs. Lyon, France; 2018.

9. Vidal A, Mengelers M, Yang S, De Saeger S, De Boevre M. Mycotoxin Biomarkers of Exposure: A Comprehensive Review. Compr Rev Food Sci Food Saf. 2018;17(5):1127– 55.

10. Ostry V, Malir F, Toman J, Grosse Y. Mycotoxins as human carcinogens—the IARC Monographs classification. Mycotoxin Res [Internet]. 2017;33(1):65–73. Available from: 10.1007/s12550-016-0265-7

11. Riley RT, Torres O, Showker JL, Zitomer NC, Matute J, Voss KA, et al. The kinetics of urinary fumonisin B1 excretion in humans consuming maize-based diets. Mol Nutr Food Res. 2012;56(9):1445–55.

12. Corrêa JAF, Orso PB, Bordin K, Hara R V, … Toxicological effects of fumonisin B1 in combination with other Fusarium toxins [Internet]. Food and Chemical …. Elsevier; 2018. Available from: https://www.sciencedirect.com/science/article/pii/S0278691518306872

13. Bakker MG, Brown DW, Kelly AC, Kim HS, … Fusarium mycotoxins: a trans-disciplinary overview. Can J … [Internet]. 2018; Available from: https://www.tandfonline.com/doi/abs/10.1080/07060661.2018.1433720

14. Chilaka CA, De Boevre M, Atanda OO, De Saeger S. The status of fusarium mycotoxins in sub-Saharan Africa: A review of emerging trends and post-harvest mitigation strategies towards food control. Vol. 9, Toxins. MDPI AG; 2017.

15. Chilaka CA, Obidiegwu JE, Chilaka AC, Atanda OO, Mally A. Mycotoxin Regulatory Status in Africa: A Decade of Weak Institutional Efforts. Toxins (Basel). 2022;14(7):1– 20.

16. Heyndrickx E, Sioen I, Huybrechts B, Callebaut A, De Henauw S, De Saeger S. Human biomonitoring of multiple mycotoxins in the Belgian population: Results of the BIOMYCO study. Environ Int. 2015;84((2015)):82–9.

17. Knutsen H-K, Barregård L, Bignami M, Brüschweiler B, Ceccatelli S, Cottrill B, et al. Appropriateness to set a group health-based guidance value for fumonisins and their modified forms. EFSA J. 2018;16(2):75pp.

18. Torres O, Matute J, Gelineau-van Waes J, Maddox JR, Gregory SG, Ashley-Koch AE, et al. Urinary fumonisin B 1 and estimated fumonisin intake in women from high-and low-exposure communities in G uatemala. Mol Nutr Food Res. 2014;58(5):973–83.

19. Knutsen HK, Alexander J, Barregård L, Bignami M, Brüschweiler B, Ceccatelli S, et al. Risks for animal health related to the presence of zearalenone and its modified forms in feed. EFSA J [Internet]. 2017;15(7):1–123. Available from: https://efsa.onlinelibrary.wiley.com/doi/abs/10.2903/j.efsa.2017.4851

20. Rivera-Núñez Z, Barrett ES, Szamreta EA, Shapses SA, Qin B, Lin Y, et al. Urinary mycoestrogens and age and height at menarche in New Jersey girls. Environ Heal A Glob Access Sci Source. 2019;18(1):1–8.

21. Zhang S, Zhou S, Gong YY, Zhao Y, Wu Y. Human dietary and internal exposure to zearalenone based on a 24-hour duplicate diet and following morning urine study. Environ Int. 2020;142((May, 2020)):105852.

22. 22. Mruczyk K, Mizgier M, … THE OCCURRENCE OF ZEARALEONE IN FOOD AND ITS EFFECT ON FERTILITY DISORDERS [Internet]. ROK XI NUMER 1 …. przeglad.wsb.net.pl; 2018. Available from: http://www.przeglad.wsb.net.pl/uploads/1/0/3/7/10371016/pmn_38_tekst.pdf#page=415

23. EFSA ECP (EFSA P on C in the FC. Scientific opinion on the appropriateness to set a group health-based guidance value for zearalenone and its modified forms. EFSA J [Internet]. 2016;14(4):46 pp. Available from: https://efsa.onlinelibrary.wiley.com/doi/abs/10.2903/j.efsa.2016.4425

24. EFSA. Scientific opinion on the risks for animal health related to the presence of zearalenone and its modified forms in feed. EFSA J [Internet]. 2017;15(7):1–123. Available from: https://efsa.onlinelibrary.wiley.com/doi/abs/10.2903/j.efsa.2017.4851

25. Liew W-P-P, Mohd-Redzwan S. Mycotoxin: Its Impact on Gut Health and Microbiota. Front Cell Infect Microbiol. 2018;8.

26. Awuchi CG, Ondari EN, Ogbonna CU, Upadhyay AK, Baran K, Okpala COR, et al. Mycotoxins Affecting Animals, Foods, Humans, and Plants: Types, Occurrence, Toxicities, Action Mechanisms, Prevention, and Detoxification Strategies-A Revisit. Foods (Basel, Switzerland). 2021 Jun;10(6).

27. Habschied K, Kanižai Šaric G, Krstanovic V, Mastanjevic K. Mycotoxins— Biomonitoring and Human Exposure. Toxins 2021, 13, 113. s Note: MDPI stays neutral with regard to jurisdictional claims in published …; 2021.

28. Raney T, Anríquez G, Croppenstedt A, Gerosa S, Lowder SK, Matuschke I, et al. The role of women in agriculture. 2011. Report No.: No. 11-02.

29. Duncan BA. Women in Agriculture in Ghana. 2004. 101 p.

30. O’Shaughnessy BR, O’Hagan AD, Burke A, McNamara J, O’Connor S. The prevalence of farmer burnout: Systematic review and narrative synthesis. J Rural Stud [Internet]. 2022;96((2022)):282–92. Available from: https://www.sciencedirect.com/science/article/pii/S0743016722002765

31. Donham KJ, Thelin A. Agricultural medicine: Rural occupational and environmental health, safety, and prevention. John Wiley & Sons; 2016. 545 pp.

32. Silva PO, Ramalho LNZ, Oliveira CAF, Ramalho FS. Reproductive, gestational, and fetal alterations induced by dietary mycotoxins: A systematic review. Pesqui Veterinária Bras. 2024;44:e07481.

33. Arifin WN. A Web-based Sample Size Calculator for Reliability Studies. Educ Med J. 2018;10(3):10 pp.

34. Fan Y, Li J, Amin K, Yu H, Yang H, Guo Z, et al. Advances in aptamers, and application of mycotoxins detection: A review. Food Res Int. 2023;170:113022.

35. Heyndrickx E, Sioen I, Huybrechts B, Callebaut A, De Henauw S, De Saeger S. Human biomonitoring of multiple mycotoxins in the Belgian population: Results of the BIOMYCO study. Environ Int. 2015;84:82–9.

36. Zhang Z, Hu X, Zhang Q, Li P. Determination for multiple mycotoxins in agricultural products using HPLC–MS/MS via a multiple antibody immunoaffinity column. J Chromatogr B. 2016 Feb 1;1021.

37. Maruvada P, Lampe JW, Wishart DS, Barupal D, Chester DN, Dodd D, et al. Perspective: Dietary Biomarkers of Intake and Exposure-Exploration with Omics Approaches. Adv Nutr. 2020 Mar;11(2):200–15.

38. Wettersten N, Katz R, Shlipak MG, Scherzer R, Waikar SS, Ix JH, et al. Urinary Biomarkers and Kidney Outcomes: Impact of Indexing Versus Adjusting for Urinary Creatinine. Kidney Med. 2021;3(4):546–554.e1.

39. Rodríguez-Carrasco Y, Berrada H, Font G, Mañes J. Multi-mycotoxin analysis in wheat semolina using an acetonitrile-based extraction procedure and gas chromatography– tandem mass spectrometry. J Chromatogr A. 2012;1270:28–40.

40. Hickert S, Gerding J, Ncube E, Hübner F, Flett B, … A new approach using micro HPLC-MS/MS for multi-mycotoxin analysis in maize samples. Mycotoxin … [Internet]. 2015; Available from: https://link.springer.com/content/pdf/10.1007/s12550-015-0221-y.pdf

41. Schieszl T, Szabó-Fodor J, Kovács M. Determination of Fusarium mycotoxin exposure in humans based on urine samples, using One Health approach: Mini-review. Acta Agrar Kaposváriensis. 2021;25(2):53–68.

42. Szabó-Fodor J, Szeitzné-Szabó M, Bóta B, Schieszl T, Angeli C, Gambacorta L, et al. Assessment of Human Mycotoxin Exposure in Hungary by Urinary Biomarker Determination and the Uncertainties of the Exposure Calculation: A Case Study. Vol. 11, Foods. 2022.

43. Rhee MY, Kim JH, Shin SJ, Gu N, Nah DY, Hong KS, et al. Estimation of 24-hour urinary sodium excretion using spot urine samples. Nutrients. 2014;6(6):2360–75.

44. Lane C, Brown M, Dunsmuir W, Kelly J, Mangos G. Can spot urine protein/creatinine ratio replace 24 h urine protein in usual clinical nephrology? Nephrology [Internet]. 2006 Jun 1;11(3):245–9. Available from: 10.1111/j.1440-1797.2006.00564.x

45. Sallsten G, Barregard L. Variability of Urinary Creatinine in Healthy Individuals. Int J Environ Res Public Health [Internet]. 2021;18(6):3166. Available from: https://www.proquest.com/scholarly-journals/variability-urinary-creatinine-healthy/docview/2628161374/se-2?accountid=150348

46. Zhang S, Ren H-F, Du R-X, Sun W-L, Fu M-L, Zhang X-C. Global, regional, and national burden of kidney dysfunction from 1990 to 2019: a systematic analysis from the global burden of disease study 2019. BMC Public Health. 2023;23(1):1218.

47. Guo J, Liu Z, Wang P, Wu H, Fan K, Jin J, et al. Global, regional, and national burden inequality of chronic kidney disease, 1990–2021: a systematic analysis for the global burden of disease study 2021. Front Med. 2025;11.

48. Pallarés N, Carballo D, Ferrer E, Rodríguez-Carrasco Y, Berrada H. High-Throughput Determination of Major Mycotoxins with Human Health Concerns in Urine by LC-Q TOF MS and Its Application to an Exposure Study. Vol. 14, Toxins. 2022.

49. Kibugu J, Munga L, Mburu D, Maloba F, Auma JE, Grace D, et al. Dietary mycotoxins: an overview on toxicokinetics, toxicodynamics, toxicity, epidemiology, detection, and their mitigation with special emphasis on aflatoxicosis in humans and animals. Toxins (Basel). 2024;16(11):483.

50. Marin S, Ramos AJ, Sanchis V. Mycotoxins: Occurrence, toxicology, and exposure assessment. Food Chem Toxicol. 2013;60:218–37.

51. de Oliveira CAF, Franco LT, Ismail A. Biomarkers for Assessing Mycotoxin Exposure and Health Effects BT - Biomarkers in Toxicology. In: Patel VB, Preedy VR, Rajendram R, editors. Cham: Springer International Publishing; 2023. p. 243–70.

52. Eshelli M, Qader MM, Jambi EJ, Hursthouse AS, Rateb ME. Current Status and Future Opportunities of Omics Tools in Mycotoxin Research. Vol. 10, Toxins. 2018.

53. Seshadri V. The inverse Gaussian distribution: statistical theory and applications. Vol. 137. Springer Science & Business Media; 2012.

54. Castell A, Arroyo-Manzanares N, Palma-Manrique R, Campillo N, Torres C, Fenoll J, et al. Evaluation of distribution of emerging mycotoxins in human tissues: applications of dispersive liquid–liquid microextraction and liquid chromatography-mass spectrometry. Anal Bioanal Chem. 2024;416(2):449–59.

55. Zeng H-Y, Li C-Y, Yao N. Fumonisin B1: A tool for exploring the multiple functions of sphingolipids in plants. Front Plant Sci. 2020;11:600458.

56. Chen J, Wen J, Tang Y, Shi J, Mu G, Yan R, et al. Research progress on fumonisin B1 contamination and toxicity: A review. Molecules. 2021;26(17):5238.

57. Faisal Z, Lemli B, Szerencsés D, Kunsági-Máté S, Bálint M, Hetényi C, et al. Interactions of zearalenone and its reduced metabolites α-zearalenol and β-zearalenol with serum albumins: Species differences, binding sites, and thermodynamics. Mycotoxin Res. 2018;34:269–78.

58. Gajęcka M, Tarasiuk M, Zielonka Ł, Dąbrowski M, Gajęcki M. Risk assessment for changes in the metabolic profile and body weights of pre-pubertal gilts during long-term monotonic exposure to low doses of zearalenone (ZEN). Res Vet Sci. 2016;109:169–80.

59. Faulk C, Barks A, Sánchez BN, Zhang Z, Anderson OS, Peterson KE, et al. Perinatal lead (Pb) exposure results in sex-specific effects on food intake, fat, weight, and insulin response across the murine life-course. PLoS One. 2014;9(8):e104273.

60. Fitzpatrick J, Schoeny R, Gallagher K, Deener K, Dockins C, Firestone M, et al. US Environmental Protection Agency’s framework for human health risk assessment to inform decision making. Int J Risk Assess Manag. 2017;20(1–3):3–20.

61. Gajecka M, Rybarczyk L, Zwierzchowski W, Jakimiuk E, Zielonka L, Obremski K, et al. Risk assessment for changes in the metabolic profile and body weights of pre-pubertal gilts during long-term monotonic exposure to low doses of zearalenone (ZEN). Res Vet Sci. 2016;109(6):169–80.

62. Mulisa G, Pero-Gascon R, McCormack V, Bisanz JE, Talukdar FR, Abebe T, et al. Multiple mycotoxin exposure assessment through human biomonitoring in an esophageal cancer case-control study in the Arsi-Bale districts of Oromia region of Ethiopia. Int J Hyg Environ Health. 2025;263:114466.

63. Strachan CR. Mycotoxins and their impact on animal health. In: World Mycotoxin Forum. 2023. p. 4.

64. De Santis B, Debegnach F, Toscano P, Crisci A, Battilani P, Brera C. Overall Exposure of European Adult Population to Mycotoxins by Statistically Modelled Biomonitoring Data. Toxins (Basel). 2021 Oct;13(10).

65. Hassan HF, Zgheib K, Iskandar CF, Chalak A, Alwan N, Abiad MG. Exposure to mycotoxins from the consumption of corn-based breakfast cereals in the United Arab Emirates. Sci Rep. 2024;14(1):25761.

66. Papageorgiou M, Wells L, Williams C, White KLM, De Santis B, Liu Y, et al. Occurrence of deoxynivalenol in an elderly cohort in the UK: a biomonitoring approach. Food Addit Contam Part A. 2018;35(10):2032–44.

67. Gong YY, Watson S, Routledge MN. Aflatoxin Exposure and Associated Human Health Effects, a Review of Epidemiological Studies. Food Saf. 2016;4(1):14–27.

68. Nahle S, El Khoury A, Atoui A. Current status on the molecular biology of zearalenone: its biosynthesis and molecular detection of zearalenone producing Fusarium species. Eur J Plant Pathol. 2021;159(2):247–58.

69. Yang S, Zhang H, Zhang J, Li Y, Jin Y, Zhang S, et al. Deglucosylation of zearalenone-14-glucoside in animals and human liver leads to underestimation of exposure to zearalenone in humans. Arch Toxicol. 2018;92(9):2779–91.

70. Faisal Z, Lemli B, Szerencsés D, Kunsági-Máté S, Bálint M, Hetényi C, et al. Interactions of zearalenone and its reduced metabolites α-zearalenol and β-zearalenol with serum albumins: species differences, binding sites, and thermodynamics. Mycotoxin Res. 2018;34(4):269–78.

